# Integrating Glaucoma Endophenotypes Improves Polygenic Risk Prediction for Primary Open-Angle Glaucoma Across Ancestries

**DOI:** 10.1101/2025.03.27.25324762

**Authors:** Xiaoyi Raymond Gao

## Abstract

Primary open-angle glaucoma (POAG) is a leading cause of irreversible blindness, and early detection is crucial. Existing polygenic risk scores (PRSs) rely on single-trait models that lack the complementary genetic information captured by glaucoma endophenotypes. We developed a multi-trait polygenic probability risk score (PPRS) framework integrating trait-specific PRSs to improve prediction across ancestries. We evaluated the PPRS in the UK Biobank (n=324,713, European ancestry) and the Mexican American Glaucoma Genetic Study (MAGGS, n=4,549, Latino ancestry). Using SBayesRC with > 7 million variants and 96 functional annotations, we constructed PRSs for POAG, intraocular pressure (IOP), vertical cup-to-disc ratio (VCDR), and retinal nerve fiber layer thickness. Our PPRS combines these trait-specific PRSs via penalized logistic regression, preserving each trait’s contribution. The PPRS achieved AUC values of 0.814 in Europeans and 0.802 in Latinos. Individuals in the highest PPRS decile had 74.4-fold (Europeans) and 49.3-fold (Latinos) higher odds of POAG than those in the lowest decile, with the top quintile capturing 65.7% and 62.2% of cases, respectively. Component trait contributions differed by ancestry: IOP PRS was strongest in Europeans (OR=1.63, *P* = 5.37 × 10^-89^), whereas VCDR PRS dominated in Latinos (OR=1.64, *P* = 2.04 × 10^-11^). Our multi-trait PPRS framework improves POAG prediction across ancestries and reveals that distinct endophenotypes drive genetic risk in different populations, supporting ancestry-informed risk stratification for targeted screening and earlier intervention. By decomposing polygenic risk into trait-specific components, this framework enables ancestry-informed glaucoma screening strategies that could facilitate earlier detection and personalized clinical management.

## Introduction

Glaucoma is the leading cause of irreversible blindness^1, 2^, affecting about 80 million individuals world-wide^3, 4^, with primary open-angle glaucoma (POAG) constituting approximately 90% of glaucoma cases in North America^4, 5^. Despite available treatments to slow disease progression, the irreversible nature of glaucoma-induced vision loss underscores the critical importance of early detection and intervention. While traditional risk assessment relies primarily on clinical measurements and family history, recent advances in genomics offer promising opportunities for enhanced risk stratification.

Polygenic risk scores (PRSs) have emerged as powerful tools for quantifying genetic predisposition to complex diseases^6–8^. However, their application to POAG has faced several key limitations. First, previous studies utilizing single-trait PRSs have achieved limited stratification ability, with only about 50% of cases being identified in the top PRS quintile in a recent report^9^. Second, existing approaches have limited capacity to capture the inherent complexity of POAG’s pathophysiology, which involves multiple quantitative endophenotypes^1, 10^, including intraocular pressure (IOP), vertical cup-to-disc ratio (VCDR), and retinal nerve fiber layer (RNFL) thickness. The multi-trait analysis of genome-wide association studies (MTAG) approach can increase power to detect associations by jointly analyzing genetically correlated traits; however, it assumes that all SNPs share the same variance-covariance matrix across traits, potentially leading to biased effect size estimates and an increased rate of false positives^11^. Third, most POAG PRS prediction studies have employed limited genomic coverage (< 1M markers) and have lacked integration of functional annotations. These limitations highlight the necessity for more sophisticated models that can capture the multifaceted genetic underpinnings of POAG.

Here, we present a multi-trait polygenic probability risk score (PPRS) approach that addresses these limitations through three key innovations: 1) integrating multiple disease-relevant quantitative traits into PRS modeling while preserving the individual contributions of each trait to enable interpretable, ancestry-specific risk assessment; 2) incorporating extensive genomic coverage (> 7 million variants) and functional annotations; 3) validating across diverse ancestral populations. We hypothesized that this comprehensive approach would significantly improve POAG risk stratification across diverse populations compared to existing methods. To test this hypothesis, we evaluated our model in two independent cohorts: the UK Biobank (UKB) for European ancestry individuals and the Mexican American Glaucoma Genetic Study (MAGGS) for Latino participants.

## Materials and Methods

### Study Design and Populations

We conducted a two-stage study: first, deriving genome-wide association study (GWAS) summary statistics from previously published data and our own GWAS analyses (details provided below); second, constructing and evaluating PRSs in a subset of the UKB cohort (n = 324,713, POAG cases: 2,168) and the MAGGS cohort (n = 4,549, POAG cases: 275), both independent of the GWAS discovery samples. UKB was approved by the North West Multi-Center Research Ethics Committee^12, 13^ and MAGGS was approved by the institutional review board at the University of Illinois at Chicago^14^. All participants provided written informed consent. The study adhered to the tenets of the Declaration of Helsinki. We obtained fully de-identified data to ensure participant confidentiality. We utilized the UKB and MAGGS datasets because both include genome-wide genotyping and detailed ophthalmic phenotypes suitable for glaucoma risk modeling. The UKB provided a large cohort enabling development and benchmarking of predictive models in participants of European ancestry, while MAGGS offered an independent Latino cohort for cross-ancestry evaluation.

### UKB dataset

The UKB is an ongoing, large-scale prospective cohort study with over 500,000 adult participants aged 40 to 69 at enrollment (2006–2010), registered with the National Health Service in the United Kingdom^12, 13^. The study collected medical information and DNA samples, including ophthalmologic data for a subset of approximately 118,000 participants. Our analysis focused on participants of European ancestry. Genotyping was performed using the UK BiLEVE Axiom Array or the UKB Axiom Array, with imputation based on reference panels from the 1000 Genomes Project (1KGP), UK10K, and the Haplotype Reference Consortium^15^. Variants with an imputation INFO score less than 0.3 and minor allele frequency less than 0.01 were excluded, resulting in approximately 10.3 million variants for downstream analysis. For individuals with IOP measurements, VCDR values, and RNFL data, we used previously published IOP GWAS results^16^, VCDR summary statistics^17^, and conducted a GWAS of RNFL to derive summary statistics. For those without ophthalmic exams (n = 324,713, unrelated, independent of the IOP, VCDR, and RNFL GWAS samples)^18^, we utilized their data for PRS construction and association testing with POAG. Importantly, individuals contributing to the IOP, VCDR, and RNFL GWAS were distinct from those used as the target sample for PRS testing, ensuring no sample overlap. POAG (H40.1) cases were identified using the International Classification of Diseases Tenth Revision (ICD-10) codes. Controls were identified as those who did not have glaucoma.

### MAGGS dataset

MAGGS samples were sourced from the Los Angeles Latino Eye Study (LALES)^19^, a population-based epidemiologic study examining visual impairment and ocular diseases in 6,357 Latino individuals aged 40 or older in Los Angeles County, California. The presence of POAG was determined by agreement among three glaucoma specialists using clinical data, including the presence of open angles, characteristic visual field abnormalities, and optic disc damage in at least one eye. MAGGS and LALES genotyped 4,996 Latino individuals using either the Illumina OmniExpress BeadChip or the Illumina Hispanic/SOL BeadChip^14, 20^. Imputation was performed using 1KGP reference panel, excluding variants with a Mach^21^ Rsq less than 0.3 and minor allele frequency less than 0.01, resulting in approximately 9.9 million imputed variants. Our analysis included 4,549 participants (4,108 unrelated and 441 related individuals) from this dataset.

### GWAS summary statistics

We used previously published GWAS summary statistics for POAG, IOP, and vertical cup-to-disc ratio (VCDR), as well as our own GWAS results for RNFL. We primarily selected publicly available GWAS summary statistics with the largest available sample sizes at the time of analysis. When suitable public datasets were unavailable, we conducted in-house GWAS to generate summary statistics. This strategy ensured the use of the most comprehensive and up-to-date data for each phenotype. The details are as follows:

1. POAG: We used GWAS results from Gharahkhani et al. (2021), which performed a multi-ethnic meta-analysis using the International Glaucoma Genetics Consortium (IGGC) and UKB datasets, comprising 383,500 individuals of European, Asian, and African descent^22^. For building PRSs for UKB participants, we used results from IGGC only, excluding UKB individuals, resulting in a cross-ancestry meta-analysis with 192,702 individuals suitable for UKB PRS construction.
2. IOP: We used GWAS results from Gao *et al.* (2018), including 115,486 UKB participants of European ancestry^16^.
3. VCDR: We used GWAS summary statistics from Han et al. (2021), involving 111,724 individuals of European and Asian descent from UKB, the Canadian Longitudinal Study on Aging (CLSA), and IGGC^17^. VCDR measurements were obtained from human graders and deep learning models applied to retinal fundus images.
4. RNFL: We conducted a GWAS with 52,902 European ancestry UKB participants using the REGENIE^23^ software and RNFL thickness values derived from optical coherence tomography images^24^. We excluded outliers and low-quality images (image quality score less than 45). We averaged RNFL values between the left and right eyes and applied rank-based inverse normalization. We adjusted for age, sex, diabetes, spherical equivalent, and the first 10 principal components of genetic ancestry, as these factors are known to influence RNFL thickness or serve as potential confounders in genetic association studies. Supplementary Figure 1 presents a Manhattan plot of the genome-wide -log10(*P* values) for our RNFL GWAS analysis.

These summary statistics provided the foundation for constructing PRSs in our analysis, tailored to different phenotypes relevant to POAG risk.

### PRS construction

We employed three different approaches to construct PRSs:

First, we implemented a traditional clumping and thresholding (C+T) based PRS^25^ using PLINK^26, 27^ to construct the POAG PRS, denoted as PRS-POAG-C+T. Independent single nucleotide polymorphisms (SNPs) were identified using linkage disequilibrium (LD)-based clumping (r² < 0.3) and selected based on a p-value threshold of *P* < 5 × 10⁻⁵.

Second, we constructed enhanced PRSs using the SBayesRC^28^ method, which leverages functional genomic annotations and extensive SNP coverage to improve polygenic risk prediction for complex traits or diseases. By integrating 96 functional annotations, including regulatory elements, chromatin states, and transcription factor binding sites, and analyzing over seven million SNPs, we sought to substantially improve PRS accuracy. Using SBayesRC, we constructed PRSs for POAG (PRS-POAG), IOP (PRS-IOP), VCDR (PRS-VCDR), and RNFL thickness (PRS-RNFL).

Third, we constructed a MTAG-derived PRS for POAG to enable direct comparison. We utilized summary statistics for 2,763 uncorrelated SNPs (identified through LD clumping at r² = 0.1 and a significance threshold of *P* < 0.001) from Craig *et al.*’s (2020) MTAG analysis^29^. The MTAG-derived PRS was calculated using the formula: PRS-POAG-MTAG(i) = ∑ β_j_ G_ij_, where β_j_ represents the regression coefficient for SNP_j_, and G_ij_ indicates the risk allele dosage for SNP_j_ in individual i. Since the MTAG summary statistics were generated using UKB data, we avoided applying the MTAG-derived PRS to UKB participants to prevent data leakage. Consequently, this PRS was exclusively applied to MAGGS participants.

### Statistical analysis

For testing the association between the PRSs and POAG, we carried out traditional logistic regression analyses, adjusting for age and sex. All constructed PRSs were standardized to have a mean of zero and a variance of one. To check potential multicollinearity among PRSs, we used two complementary approaches. First, we computed the variance inflation factor (VIF) for each predictor, noting that VIF values less than 5 typically indicate low correlation^30^. Second, we employed XGBoost^31^, a tree-based method known to be less sensitive to collinearity, and examined feature importances via SHapley Additive exPlanations (SHAP) values^32^, following our previous methodology^33^.

For assessing the predictive ability of the PRSs for POAG, we used penalized (L2) logistic regression with stratified 10-fold cross-validation performed separately in the UKB and MAGGS datasets. We defined the polygenic probability risk score (PPRS) as the predicted probability output from an L2-penalized logistic regression model (scikit-learn LogisticRegression with penalty = “l2” ^34^), which integrates four PRSs (PRS-POAG, PRS-IOP, PRS-VCDR, and PRS-RNFL) together with age and sex as covariates. The model takes these inputs and outputs a probability of glaucoma for each individual, which we term the PPRS. This formulation differentiates the PPRS from traditional single-trait PRSs and the MTAG-derived PRS. For MAGGS related samples, predictive models were developed using MAGGS unrelated samples and subsequently applied to the related individuals to derive predicted probabilities. We used the area under the receiver operating characteristic curve (AUC) to quantify the predictive ability of the PPRS on POAG. Pairwise comparisons of AUC between different models were performed using Delong’s test^35^. For risk stratification, we divided the PPRS into deciles and compared the odds of POAG among study participants in higher PPRS deciles versus those in the lowest PPRS decile in both the UKB and MAGGS datasets. A statistical significance threshold of *P* < 0.05 was used for all analyses. All analyses were performed using R (v4.4; including the ggplot2 package for plotting), SAS 9.4 (SAS, Inc., Cary, NC, for non-penalized logistic regression), and Python (v3.12; scikit-learn for penalized logistic regression, XGBoost for gradient boosting models, and SHAP for model interpretability).

## Results

Table 1 summarizes the characteristics of our study samples from UKB and MAGGS. The UKB cohort comprised 324,713 individuals with 2,168 glaucoma cases and 322,545 controls. The mean (standard deviation [SD]) age of glaucoma and non-glaucoma was 63.0 (5.7) and 57.1 (8.0) years, respectively. Females consisted of 48.4% of glaucoma cases and 54.3% of controls. The MAGGS dataset included 4,549 participants with 275 glaucoma cases and 4,274 controls. The mean (SD) age of glaucoma and non-glaucoma was 66.4 (11.1) and 56.1 (10.2), respectively. Females constituted 52.4% of glaucoma cases and 59.2% of controls.

**Table 1.** Sample Characteristics.

|  | UK Biobank (n = 324,713) |  | MAGGS (n = 4,549) |  |
| --- | --- | --- | --- | --- |
|  | Controls (322,545) | Cases (2,168) | Controls (4,274) | Cases (275) |
| Age (yrs, SD) | 57.1 (8.0) | 63.0 (5.7) | 56.1 (10.2) | 66.4 (11.1) |
| Female (%) | 54.3 | 48.4 | 59.2 | 52.4 |

Abbreviation: MAGGS, Mexican American Glaucoma Genetic Study; SD: standard deviation.

Table 2 presents the single PRS association results (logistic regression analyses), adjusted for age and sex, among UKB and MAGGS participants. In the UKB cohort (Table 2a), the traditional clumping and thresholding (C+T) PRS for POAG (PRS-POAG-C+T) yielded an odds ratio (OR) of 1.71 (95% confidence interval [CI]: 1.64–1.79; *P* = 8.71 × 10^−141^). The SBayesRC-derived PRS for POAG (PRS-POAG, which integrates functional annotation information) showed a stronger association with an OR of 2.14 (95% CI: 2.05–2.24; *P* = 8.54 × 10^−269^). The SBayesRC-derived PRSs for IOP (PRS-IOP), VCDR (PRS-VCDR), and RNFL thickness (PRS-RNFL) also demonstrated significant associations with POAG. Similarly, in the MAGGS cohort (Table 2b), the PRS-POAG-MTAG had an OR of 1.44 (95% CI: 1.24–1.60; *P* = 1.17 × 10^−7^). PRS-VCDR showed the highest OR of 1.90 (95% CI: 1.66-2.17; *P* = 1.39 × 10^−20^). The PRS-POAG-C+T, PRS-IOP, PRS-POAG, and PRS-RNFL in MAGGS participants also exhibited significant associations with POAG.

**Table 2.**
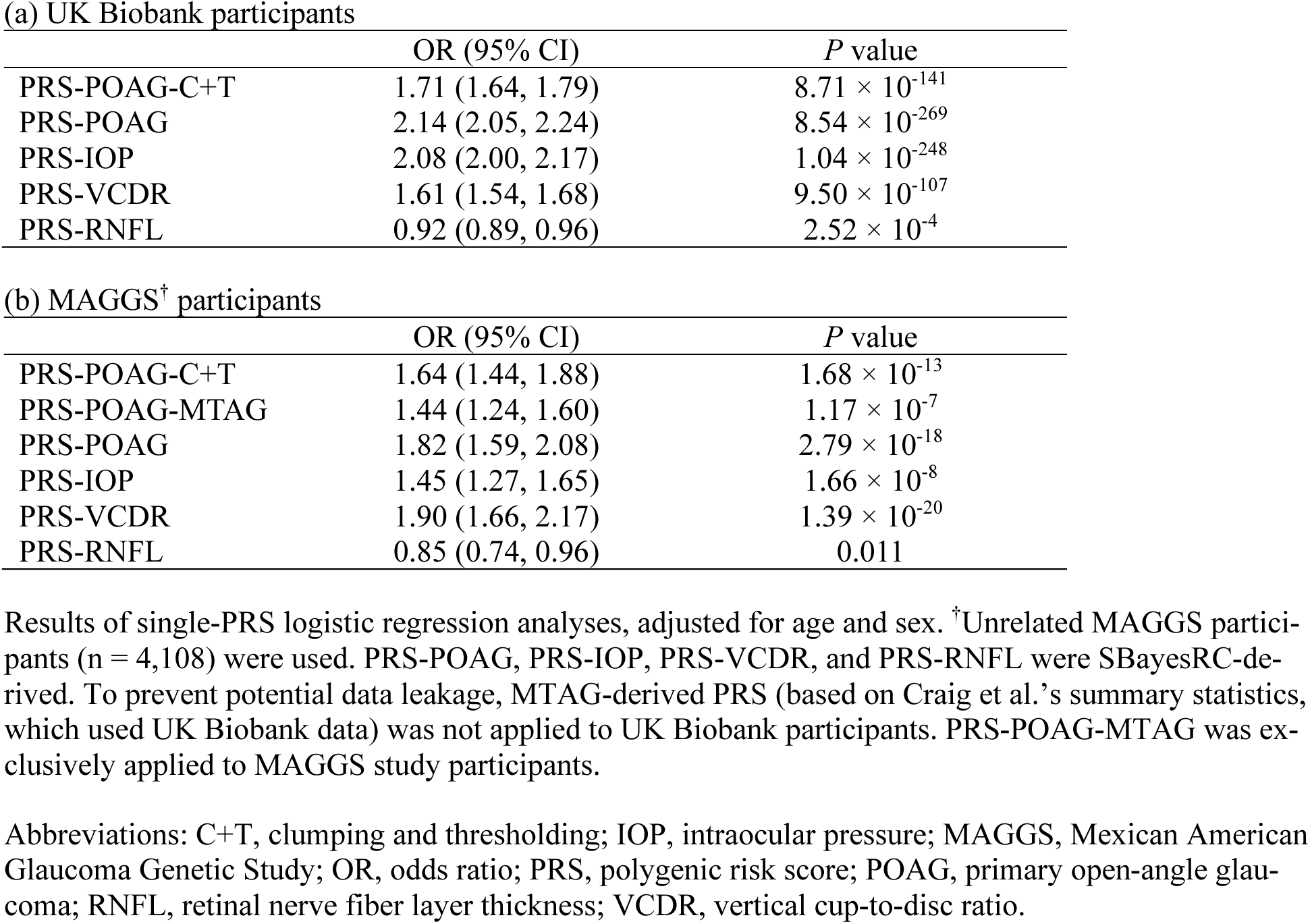
Single-PRS Logistic Regression Results.

Table 3 displays the results of the multiple PRS logistic regression analyses, assessing the combined association of PRSs with POAG and their relative importance in each cohort. In the UKB participants, PRS-POAG had an OR of 1.59 (*P* = 1.23 × 10^−74^), PRS-IOP showed the strongest association with an OR of 1.63 (*P* = 5.37 × 10^−89^), PRS-VCDR had an OR of 1.28 (*P* = 1.99 × 10^−27^), and PRS-RNFL exhibited an OR of 0.94 (*P* = 0.002), indicating a slight inverse association (larger PRS-RNFL indicated thicker RNFL and hence is protective). The OR for a simultaneous one-SD-unit-increase in all four standardized PRS variables was 3.11 (95% CI: 2.89-3.34). In the MAGGS cohort, PRS-POAG had an OR of 1.40 (*P* = 7.63 × 10^−5^), PRS-IOP an OR of 1.15 (*P* = 0.07), PRS-VCDR showed the strongest association with an OR of 1.64 (*P* = 2.04 × 10^−11^), and PRS-RNFL had an OR of 0.85 (*P* = 0.016), also suggesting an inverse relationship. The OR for a simultaneous one-SD-unit-increase in all four standardized PRS variables was 2.25 (95% CI: 1.79-2.82). These results highlight ancestry-specific differences in genetic contributions to POAG risk, with PRS-IOP contributing most significantly in the UKB dataset (European ancestry), whereas PRS-VCDR had the strongest contribution in the MAGGS dataset (Latino ancestry). We further evaluated potential collinearity among these PRSs using VIF (Supplementary Table 1) and XGBoost with SHAP-based feature importance (Supplementary Figure 2). Both methods indicated no collinearity issues, underscoring the robustness of our logistic regression analyses.

**Table 3.** Multiple Logistic Regression Results.

|  | UK Biobank |  | MAGGS<sup>†</sup> |  |
| --- | --- | --- | --- | --- |
|  | OR (95% CI) | *P* value | OR (95% CI) | *P* value |
| PRS-POAG | 1.59 (1.51, 1.67) | $1.23 \times 10^{-74}$ | 1.40 (1.18, 1.65) | $7.63 \times 10^{-5}$ |
| PRS-IOP | 1.63 (1.55, 1.71) | $5.37 \times 10^{-89}$ | 1.15 (0.99, 1.35) | 0.07 |
| PRS-VCDR | 1.28 (1.23, 1.34) | $1.99 \times 10^{-27}$ | 1.64 (1.42, 1.90) | $2.04 \times 10^{-11}$ |
| PRS-RNFL | 0.94 (0.90, 0.98) | 0.002 | 0.85 (0.74, 0.97) | 0.016 |

Results of multiple logistic regression analyses, adjusted for age and sex, assessing the combined association of polygenic risk scores (PRSs) with primary open-angle glaucoma. <sup>†</sup>Unrelated MAGGS participants (n = 4,108) were used. PRS-POAG, PRS-IOP, PRS-VCDR, and PRS-RNFL were SBayesRC-derived.
Abbreviations: IOP, intraocular pressure; MAGGS, Mexican American Glaucoma Genetic Study; OR, odds ratio; PRS, polygenic risk score; POAG, primary open-angle glaucoma; RNFL, retinal nerve fiber layer thickness; VCDR, vertical cup-to-disc ratio.

We then examined the discriminatory ability of the PRSs (PRS-POAG, PRS-IOP, PRS-VCDR, and PRS-RNFL) for predicting POAG using penalized (L2) logistic regression with stratified 10-fold cross-validation and calculated the area under the receiver operating characteristic curve (AUC). Figure 1 illustrates these results. In UKB participants (Figure 1A), the baseline model including only age and sex had an AUC of 0.721 (95% CI: 0.711-0.730). Adding PRS-POAG-C+T increased the AUC to 0.761 (95% CI: 0.752-0.771), while incorporating the SBayesRC-derived PRS-POAG improved it to 0.790 (95% CI: 0.781-0.799), a 3% increase over the C+T approach (*P* = 1.13 × 10^−29^). Incorporating all four PRSs (PRS-POAG, PRS-IOP, PRS-VCDR, and PRS-RNFL) with adjustment for age and sex resulted in an AUC of 0.814 (95% CI: 0.806–0.823), representing a 9.3% improvement over the baseline model (*P* = 1.15 × 10^−112^) and a 2.1% improvement over the SBayesRC-derived PRS-POAG (*P* = 6.25 × 10^−28^). In MAGGS participants (Figure 1B), the baseline AUC was 0.753 (95% CI: 0.724-0.782), which increased to 0.761 (95% CI: 0.732-0.789) with PRS-POAG-MTAG, to 0.773 (95% CI: 0.744-0.802) with PRS-POAG-C+T, and to 0.782 (95% CI: 0.754-0.810) with PRS-POAG. The full model with all four PRSs (PRS-POAG, PRS-IOP, PRS-VCDR, and PRS-RNFL) achieved an AUC of 0.802 (95% CI: 0.775–0.828), providing a 5% enhancement over the baseline model (*P* = 5.25 × 10^−6^) and a 2% enhancement over the SBayesRC-derived PRS-POAG (*P* = 9.92 × 10^−4^). These findings indicate that incorporating multiple glaucoma-related PRSs significantly improves the prediction of POAG in both cohorts.

**Figure 1.**
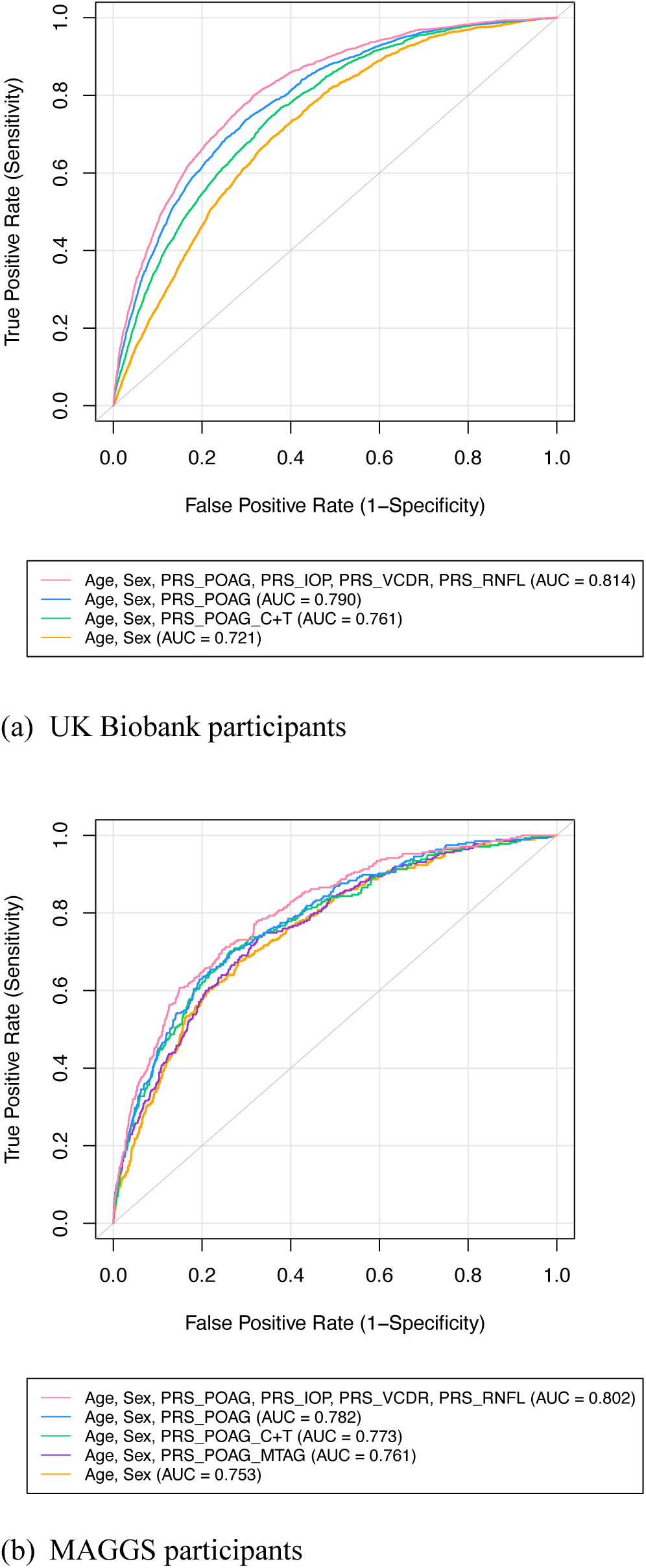
Receiver Operating Characteristic Curves Predicting POAG Using Polygenic Risk Scores. Receiver operating characteristic curves illustrating the predictive performance of polygenic risk scores (PRSs) for primary open-angle glaucoma (POAG) in (A) the UK Biobank cohort and (B) the Mexican American Glaucoma Genetic Study (MAGGS) cohort. In both cohorts, models incorporating age, sex, and multi-trait PRSs significantly improved the area under the curve (AUC) compared to the baseline models (age and sex only) and single-PRS models. Analysis was performed using penalized (L2) logistic regression with stratified 10-fold cross-validation. Abbreviations: IOP, intraocular pressure; RNFL, retinal nerve fiber layer; VCDR, vertical cup-to-disc ratio.

We further evaluated POAG risk stratification using polygenic probability risk score (PPRS), integrating the effect of multiple PRSs of POAG, IOP, VCDR, and RNFL (with adjustment for age and sex), from our penalized 10-fold cross-validation logistic regression models. Analysis of PPRS revealed strong risk stratification capabilities (Figure 2). In the UKB cohort (Figure 2A), a clear trend emerged with increasing ORs across higher PPRS deciles. Individuals in the highest decile had 74.35-fold increased odds of POAG (95% CI: 43.87–126.02; *P* = 1.28 × 10^−57^) compared with those in the lowest decile, and 12.82-fold increased odds (95% CI: 10.22–16.09; *P* = 1.17 × 10^−107^) compared with those in the 5^th^ decile. Similarly, in the MAGGS cohort (Figure 2B), participants in the highest decile demonstrated 49.34-fold increased odds of POAG (95% CI: 15.54–156.62; *P* = 3.69 × 10^−11^) compared with the lowest decile and 9.61-fold increased odds (95% CI: 5.50–16.77; *P* = 1.72 × 10^−15^) compared with the 5^th^ decile. Most POAG cases (46.54% in UKB and 40.73% in MAGGS) fell within the highest decile category of PPRS (Supplementary Table 2). Taken together, these findings underscore the strong relationship between PPRS extreme deciles and POAG risk across UKB and MAGGS, highlighting the potential utility of the PPRS for POAG risk stratification.

**Figure 2.**
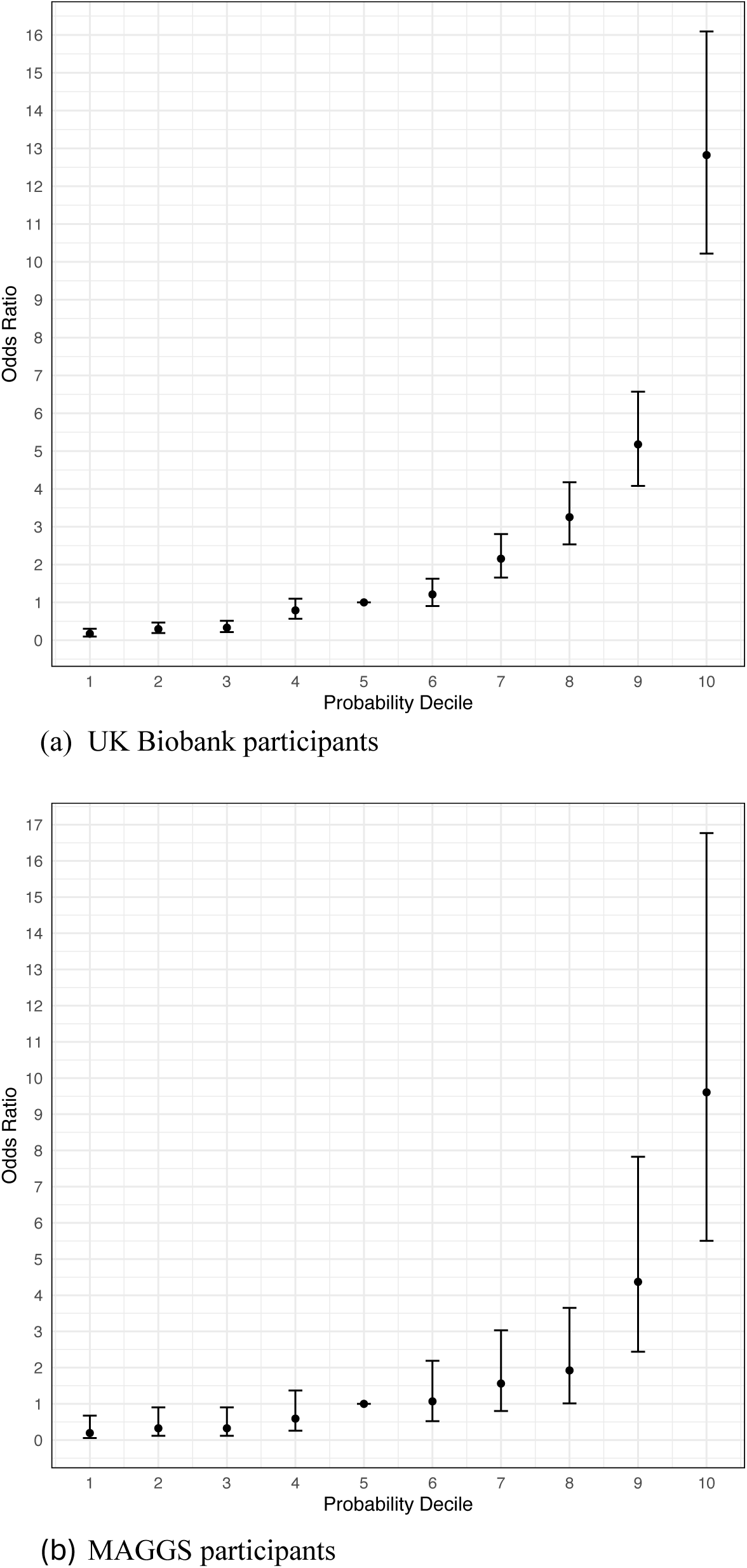
Association Between Polygenic Probability Risk Score Deciles and POAG Risk. Odds ratios (ORs) for primary open-angle glaucoma (POAG) across deciles of the polygenic probability risk score (PPRS) in (A) the UK Biobank cohort and (B) the Mexican American Glaucoma Genetic Study (MAGGS) cohort. The fifth decile (Decile 5) serves as the reference group. Error bars represent 95% confidence intervals (CIs). A clear trend of increasing POAG risk with higher PPRS deciles demonstrates strong risk stratification capabilities of PPRS in both ancestral cohorts.

## Discussion

Our study demonstrates that integrating multiple glaucoma-related traits with functional annotations substantially improves the polygenic prediction for POAG across diverse ancestries. Three key findings emerge from our analysis: First, our multi-modal PPRS approach achieved higher predictive accuracy, with AUC values of 0.814 (95% CI: 0.806–0.823) in Europeans and 0.802 (95% CI: 0.775–0.828) in Latinos, compared to our POAG single-trait models. This improvement likely reflects the complex genetic architecture of POAG, where multiple biological pathways contribute to disease development. Second, we identified ancestry-specific differences in the relative contributions of component PRSs. In Europeans, IOP showed the strongest association, whereas VCDR was most strongly associated in Latinos. This finding underscores the importance of population-specific risk assessment and suggests distinct genetic mechanisms underlying POAG across ancestral groups. Third, the strong risk stratification achieved by our PPRS, demonstrating over 74.4-fold and 49.3-fold risk differences between extreme deciles in Europeans and Latinos, respectively, suggests significant clinical utility. Importantly, unlike approaches that collapse multiple traits into a single combined score (e.g., via multi-trait analysis of GWAS), our framework preserves trait-specific PRS contributions, enabling the identification of ancestry-specific differences in the genetic architecture underlying POAG risk.

One of the most direct potential clinical applications of PRS is identifying high-risk individuals. MacGregor et al. (2018), using an IOP PRS, reported an increased glaucoma risk (OR = 5.6, 95% CI: 4.1–7.6) comparing the top and bottom deciles^36^. Craig et al. (2020), employing an MTAG approach for their POAG PRS, observed a higher OR of 14.9 (95% CI: 10.7–20.9) for comparing the top and bottom deciles^29^. Gao et al. (2019) found an OR of 6.3 for their top IOP PRS quintile versus the bottom quintile comparison, with approximately 40.6% of POAG cases found in the top quintile^18^. Similarly, de Vries et al. (2025) identified about 50% of cases in their top glaucoma PRS quintile^9^. Our study substantially improves upon these results, demonstrating ORs of 74.4 (95% CI: 43.9–126.0) in Europeans and 49.3 (95% CI: 15.5–156.6) in Latinos when comparing extreme deciles of our PPRS. Additionally, we identified 65.7% and 62.2% of glaucoma cases within the top quintile of our PPRS scores for Europeans and Latinos (Supplementary Table 2), respectively, representing a substantial improvement over previous studies.

While the improvement in AUC from the best single PRS (PRS-POAG, AUC = 0.790) to the full PPRS model (AUC = 0.814) may appear modest in absolute terms (ΔAUC = 0.024), such increments are consistent with prior reports in complex disease genetics and can nonetheless have important implications in practice. In our study, the PPRS not only improved overall discrimination but also strengthened risk separation at the extremes of the distribution, with substantially elevated odds of glaucoma observed in the top decile compared with the bottom decile. Most importantly, the full PPRS model identified 12.1% more cases in the top decile than the PRS-POAG model. These tail effects are clinically relevant for identifying individuals most likely to benefit from targeted surveillance or early intervention. We therefore view the statistically significant increment in AUC as both supportive of the additive value of integrating multiple glaucoma-related PRSs and a foundation for further improvement when combined with imaging, electronic health record, or other multimodal data in future predictive models.

The observation that PRS-IOP contributed most strongly to prediction in Europeans, whereas PRS-VCDR appeared more influential in Latinos, is intriguing but should be interpreted with caution. These differences may partly reflect disparities in sample size and statistical power across discovery GWAS. Cohort-specific factors such as recruitment differences, phenotype definitions, and environmental exposures may also have influenced the relative performance of the PRSs. Nevertheless, prior studies have shown that IOP PRS significantly improves discrimination for glaucoma in Europeans^18^, and over 72% of known POAG risk loci are also associated with IOP in predominantly European datasets^22^, underscoring the key role of IOP in POAG pathogenesis in this ancestry group. At the same time, epidemiological studies report variation in POAG presentation across populations^37^: in Latinos, the majority of cases occur with IOP < 21 mmHg, with elevated IOP (>21 mmHg) observed in only ∼18% of cases. This pattern is consistent with stronger predictive value of VCDR-based genetic risk in the MAGGS cohort. The borderline non-significance of PRS-IOP in the MAGGS multiple regression model (*P* = 0.07) underscores the need for caution, as the limited sample size constrains power for precise estimates. Additional studies in larger and more diverse cohorts will be essential to further validate these ancestry-specific patterns.

Concurrent with our work, MacGregor *et al*.^38^ developed a glaucoma PRS using SBayesRC and data from over 6 million individuals, including 23andMe participants, and demonstrated strong risk stratification across five ancestry groups (European, African, East Asian, South Asian, and Admixed American/Hispanic) as well as clinical utility in predicting treatment escalation and surgical intervention. While their approach achieves impressive performance through a substantially larger discovery sample and a single combined MTAG-derived PRS, our multi-trait PPRS framework provides complementary and distinct insights. Specifically, by retaining separate trait-specific PRSs (for POAG, IOP, VCDR, and RNFL) and combining them at the prediction model level, our approach reveals that the relative genetic contributions of these component traits differ across ancestries, with IOP contributing most strongly in Europeans and VCDR in Latinos. This finding has implications for understanding the heterogeneous pathophysiology of POAG across populations and for tailoring future risk assessment strategies. Moreover, the two approaches are not mutually exclusive: incorporating our multi-trait integration framework with the larger discovery datasets used by MacGregor *et al*. could further improve prediction accuracy and interpretability in future work.

Our study has several strengths, notably the use of a comprehensive multi-trait PPRS approach that integrates multiple glaucoma-related traits and leverages functional annotations to improve prediction accuracy. By evaluating PRS performance in both European and Latino populations, we address the crucial need for inclusive and representative genetic research, especially among traditionally understudied groups^39^. However, our study also has limitations. Due to resource constraints, we included only European and Latino populations in this study. Expanding future research to additional ancestry populations would enhance generalizability and ensure broader applicability of our findings. In addition, the smaller sample size of the MAGGS Latino cohort limits the precision of effect estimates within this ancestry, underscoring the need for replication in larger datasets. Furthermore, our PRS construction was limited to common variants. Incorporating rare variants, similar to previous reports on rare variants associated with IOP^40^, warrants further investigation. Additionally, compared with recent large-scale efforts that leverage discovery samples exceeding 6 million individuals, our GWAS discovery samples are substantially smaller; however, the purpose of our study is to demonstrate the added value of a multi-trait integration framework rather than to maximize single-trait PRS performance through sample size alone. Future studies combining our multi-trait approach with larger multi-ancestry discovery datasets may yield further improvements.

It is noteworthy that the MTAG-derived PRS did not outperform the single-trait GWAS PRS in our analysis. Several factors likely contributed. First, we applied MTAG only to the MAGGS Latino samples, where the smaller cohort size may limit stable estimation. Second, we constructed PRS using the state-of-the-art SBayesRC framework, which leverages functional annotations to increase power. Finally, although the recently published large-scale POAG MTAG GWAS^41^ included >600,000 participants and could potentially offer improved power, only p-values (and not SNP effect sizes) were released, precluding its direct use for PRS derivation in this study. Notably, our multi-trait PPRS approach circumvents the limitations of MTAG by instead combining individual trait-specific PRSs at the prediction model level, which avoids MTAG’s assumption of a shared variance–covariance structure across all SNPs and enables the assessment of differential trait contributions across ancestries.

In conclusion, our multi-trait PPRS approach, combining multiple glaucoma-related traits and incorporating functional annotations, represents a significant advancement in polygenic risk prediction for POAG. By retaining trait-specific PRS contributions rather than collapsing them into a single score, our framework uniquely reveals that the genetic determinants of POAG risk differ across ancestries, with IOP-related genetic variation contributing most strongly in Europeans and VCDR-related variation in Latinos. The robust performance across different ancestral groups and substantial risk stratification capabilities suggest considerable potential for enhancing early detection and preventative strategies. By facilitating the identification of individuals at high genetic risk, our approach has significant implications for timely clinical intervention and could ultimately reduce the public health burden of POAG.

## Data Availability

All data produced in the present work are contained in the manuscript.

## Acknowledgements

We would like to thank the study participants from the UK Biobank, the Los Angeles Latino Eye Study, the Mexican American Glaucoma Genetic Study, and the staff who aided in data collection and processing. This work was supported in part by National Institutes of Health (NIH; Bethesda, MD, USA) grants P30EY032857 and R01EY022651. The content is solely the responsibility of the authors and does not necessarily represent the official views of the NIH.

## Author Contributions

X.R.G. conceived the present study, analyzed the data and wrote the manuscript.

## Data availability statement

The SNP weight data used in this study are available at https://github.com/xraygao/PPRS.

---

**Supplementary Table 1.** Variance Inflation factors for polygenic risk scores in our logistic regression models.

(a) UK Biobank
| Age | Sex | PRS-POAG | PRS-IOP | PRS-VCDR | PRS-RNFL |
| --- | --- | --- | --- | --- | --- |
| 1.002 | 1.002 | 1.347 | 1.240 | 1.112 | 1.003 |

(b) MAGGS<sup>†</sup>
| Age | Sex | PRS-POAG | PRS-IOP | PRS-VCDR | PRS-RNFL |
| --- | --- | --- | --- | --- | --- |
| 1.024 | 1.006 | 1.555 | 1.371 | 1.166 | 1.004 |

Displayed are the variance inflation factors for the polygenic risk scores (PRSs) used in our logistic regression analyses of a) UK Biobank; b) MAGGS. <sup>†</sup>Unrelated MAGGS participants were used. PRS-POAG, PRS-IOP, PRS-VCDR, and PRS-RNFL were SBayesRC-derived. These variant inflation factor values are only slightly above 1 and all remain well below the typical cutoff of 5, indicating no collinearity issues.
Abbreviations: IOP, intraocular pressure; MAGGS, Mexican American Glaucoma Genetic Study; PRS, polygenic risk score; POAG, primary open-angle glaucoma; RNFL, retinal nerve fiber layer thickness; VCDR, vertical cup-to-disc ratio.

**Supplementary Table 2.** Number of POAG Cases in Each Decile Category.

(a) UK Biobank
|  | Predicted Probability Risk Score Decile |  |  |  |  |  |  |  |  |  |
| --- | --- | --- | --- | --- | --- | --- | --- | --- | --- | --- |
|  | 1 | 2 | 3 | 4 | 5 | 6 | 7 | 8 | 9 | 10 |
|  | 14 | 24 | 27 | 64 | 81 | 98 | 174 | 262 | 415 | 1009 |
| POAG, n (%) | (0.65) | (1.11) | (1.25) | (2.95) | (3.74) | (4.52) | (8.03) | (12.08) | (19.14) | (46.54) |

(b) MAGGS
|  | Predicted Probability Risk Score Decile |  |  |  |  |  |  |  |  |  |
| --- | --- | --- | --- | --- | --- | --- | --- | --- | --- | --- |
|  | 1 | 2 | 3 | 4 | 5 | 6 | 7 | 8 | 9 | 10 |
|  | 3 | 5 | 5 | 9 | 15 | 16 | 23 | 28 | 59 | 112 |
| POAG, n (%) | (1.09) | (1.82) | (1.82) | (3.27) | (5.45) | (5.82) | (8.36) | (10.18) | (21.45) | (40.73) |

**Supplementary Figure 1.**
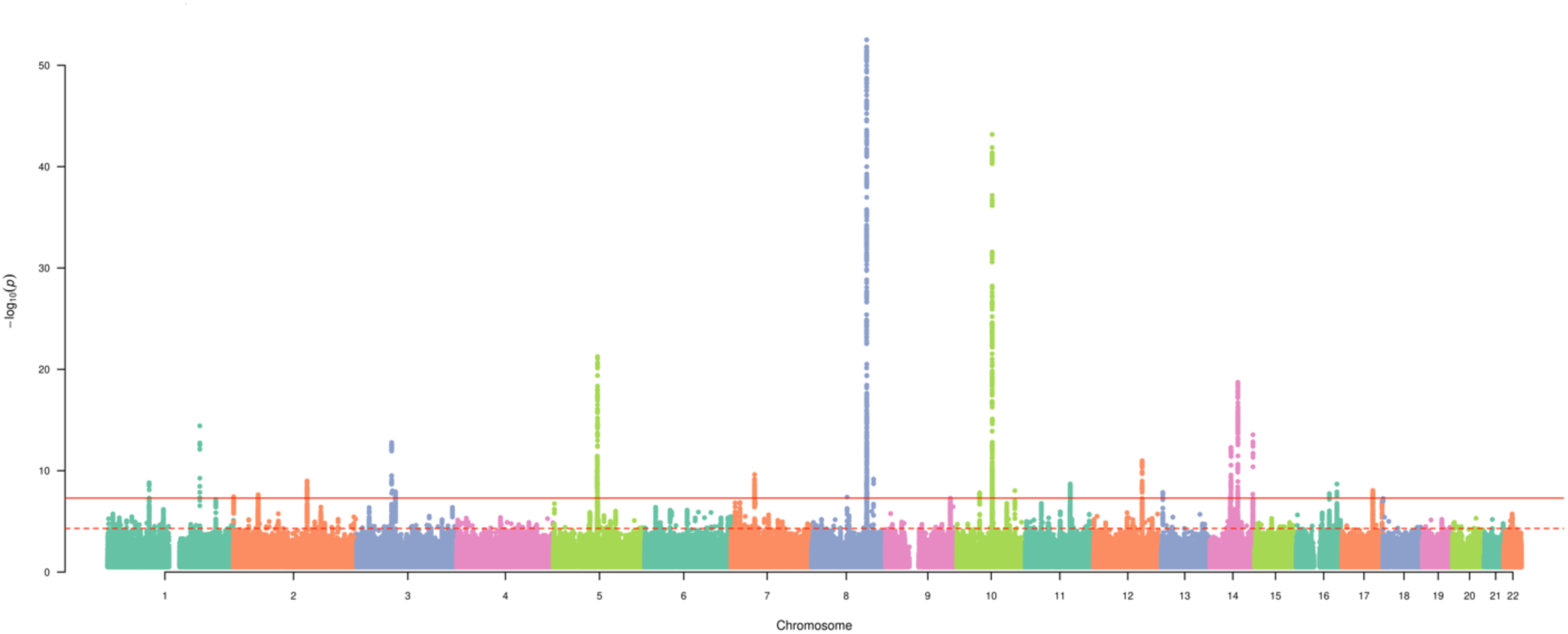
Manhattan Plot Displaying the –log10(*P* values) for the Association Between RNFL Thickness and Genome-wide Genetic Variants. Solid and dotted horizontal lines represent genome-wide significant associations (*P* < 5 ×10^-8^) and suggestive associations (*P* < 5 ×10^-5^), respectively. Genetic variants are plotted by chromosomal position.

**Supplementary Figure 2.**
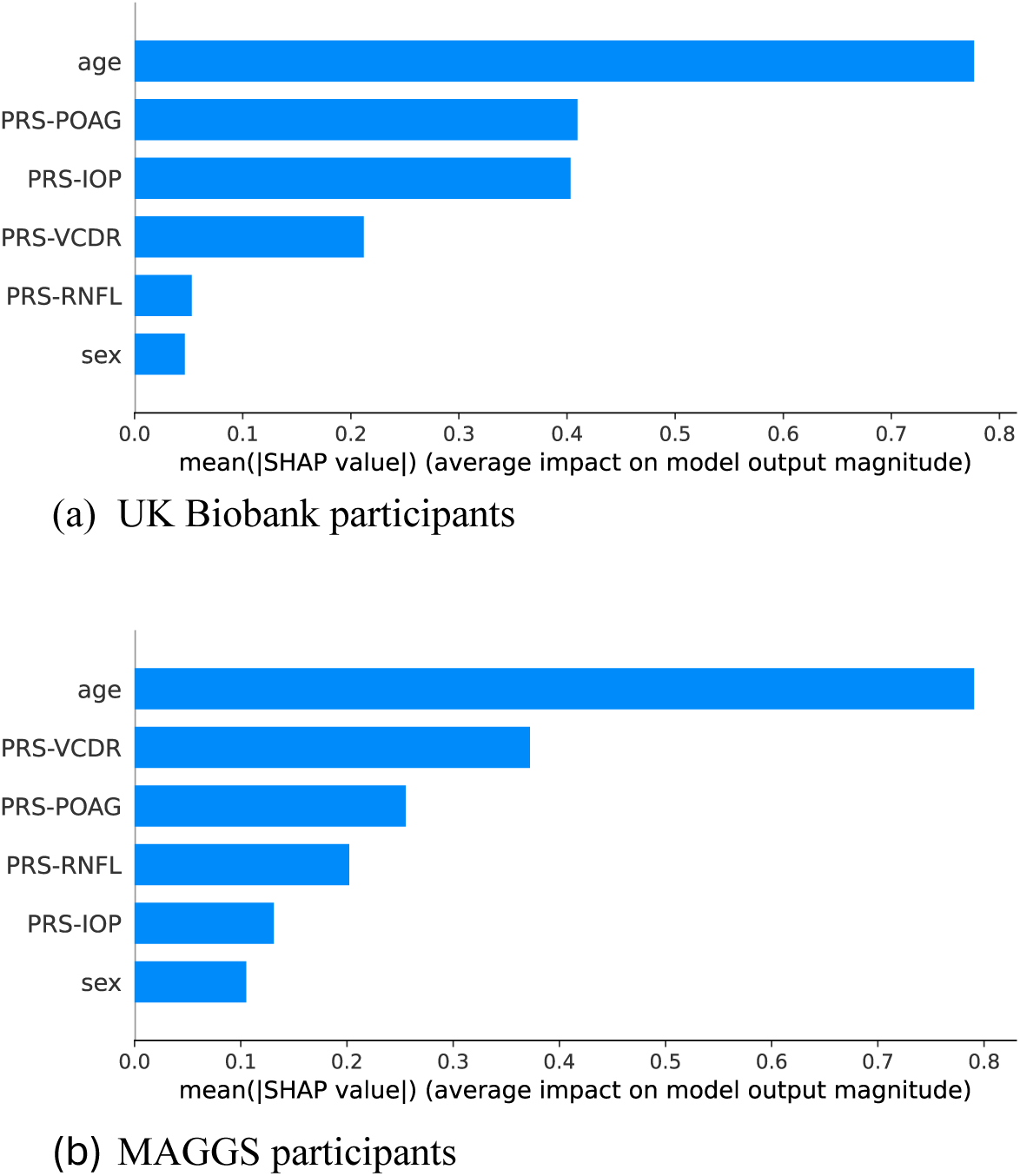
SHAP Feature Importance for XGBoost Models. Bar charts illustrating feature importance, as evaluated via SHapley Additive exPlanations (SHAP) values, for the (A) UK Biobank and (B) Mexican American Glaucoma Genetic Study (MAGGS) cohorts. The y-axis displays the ranked features, and the x-axis shows the average impact on the model’s output. All PRSs (PRS-POAG, PRS-IOP, PRS-VCDR, and PRS-RNFL) were SBayesRC-derived. In UKB participants (European ancestry), PRS-IOP and PRS-POAG both rank highly among the PRSs, whereas in MAGGS participants (Latino ancestry), PRS-VCDR shows the highest importance among the PRSs. These results align with the logistic regression findings, confirming ancestry-specific differences in genetic contributions to POAG risk. Abbreviations: IOP, intraocular pressure; RNFL, retinal nerve fiber layer; VCDR, vertical cup-to-disc ratio.

## References

1. Fan BJ, Wiggs JL. Glaucoma: genes, phenotypes, and new directions for therapy. J Clin Invest 2010;120(9):3064–72.

2. Blindness GBD, Vision Impairment C, Vision Loss Expert Group of the Global Burden of Disease S. Causes of blindness and vision impairment in 2020 and trends over 30 years, and prevalence of avoidable blindness in relation to VISION 2020: the Right to Sight: an analysis for the Global Burden of Disease Study. Lancet Glob Health 2021;9(2):e144–e60.

3. Weinreb RN, Leung CK, Crowston JG, et al. Primary open-angle glaucoma. Nat Rev Dis Primers 2016;2:16067.

4. Tham YC, Li X, Wong TY, et al. Global prevalence of glaucoma and projections of glaucoma burden through 2040: a systematic review and meta-analysis. Ophthalmology 2014;121(11):2081–90.

5. Lang GK. Ophthalmology: A Pocket Textbook Atlas, 2nd ed. Stuttgart, New Yok: Thieme 2007.

6. Purcell SM, Wray NR, Stone JL, et al. Common polygenic variation contributes to risk of schizophrenia and bipolar disorder. Nature 2009;460(7256):748–52.

7. Kachuri L, Chatterjee N, Hirbo J, et al. Principles and methods for transferring polygenic risk scores across global populations. Nat Rev Genet 2024;25(1):8–25.

8. Torkamani A, Wineinger NE, Topol EJ. The personal and clinical utility of polygenic risk scores. Nat Rev Genet 2018;19(9):581–90.

9. de Vries VA, Hanyuda A, Vergroesen JE, et al. The Clinical Usefulness of a Glaucoma Polygenic Risk Score in 4 Population-Based European Ancestry Cohorts. Ophthalmology 2025;132(2):228–37.

10. Ramdas WD, Amin N, van Koolwijk LM, et al. Genetic architecture of open angle glaucoma and related determinants. J Med Genet 2011;48(3):190–6.

11. Turley P, Walters RK, Maghzian O, et al. Multi-trait analysis of genome-wide association summary statistics using MTAG. Nat Genet 2018;50(2):229–37.

12. Sudlow C, Gallacher J, Allen N, et al. UK biobank: an open access resource for identifying the causes of a wide range of complex diseases of middle and old age. PLoS Med 2015;12(3):e1001779.

13. Allen NE, Sudlow C, Peakman T, et al. UK biobank data: come and get it. Sci Transl Med 2014;6(224):224ed4.

14. Nannini DR, Torres M, Chen YI, et al. A Genome-Wide Association Study of Vertical Cup-Disc Ratio in a Latino Population. Invest Ophthalmol Vis Sci 2017;58(1):87–95.

15. Bycroft C, Freeman C, Petkova D, et al. The UK Biobank resource with deep phenotyping and genomic data. Nature 2018;562(7726):203–9.

16. Gao XR, Huang H, Nannini DR, et al. Genome-wide association analyses identify new loci influencing intraocular pressure. Hum Mol Genet 2018;27(12):2205–13.

17. Han X, Steven K, Qassim A, et al. Automated AI labeling of optic nerve head enables insights into cross-ancestry glaucoma risk and genetic discovery in >280,000 images from UKB and CLSA. Am J Hum Genet 2021.

18. Gao XR, Huang H, Kim H. Polygenic Risk Score Is Associated With Intraocular Pressure and Improves Glaucoma Prediction in the UK Biobank Cohort. Transl Vis Sci Technol 2019;8(2):10.

19. Varma R, Paz SH, Azen SP, et al. The Los Angeles Latino Eye Study: design, methods, and baseline data. Ophthalmology 2004;111(6):1121–31.

20. Nannini D, Torres M, Chen YD, et al. African Ancestry Is Associated with Higher Intraocular Pressure in Latinos. Ophthalmology 2016;123(1):102–8.

21. Li Y, Abecasis GR. Mach 1.0: Rapid haplotype reconstruction and missing genotype inference. Am J Hum Genet 2006;S79.

22. Gharahkhani P, Jorgenson E, Hysi P, et al. Genome-wide meta-analysis identifies 127 open-angle glaucoma loci with consistent effect across ancestries. Nat Commun 2021;12(1):1258.

23. Mbatchou J, Barnard L, Backman J, et al. Computationally efficient whole-genome regression for quantitative and binary traits. Nat Genet 2021;53(7):1097–103.

24. Ko F, Foster PJ, Strouthidis NG, et al. Associations with Retinal Pigment Epithelium Thickness Measures in a Large Cohort: Results from the UK Biobank. Ophthalmology 2017;124(1):105–17.

25. Prive F, Vilhjalmsson BJ, Aschard H, Blum MGB. Making the Most of Clumping and Thresholding for Polygenic Scores. Am J Hum Genet 2019;105(6):1213–21.

26. Purcell S, Neale B, Todd-Brown K, et al. PLINK: a tool set for whole-genome association and population-based linkage analyses. Am J Hum Genet 2007;81(3):559–75.

27. Chang CC, Chow CC, Tellier LC, et al. Second-generation PLINK: rising to the challenge of larger and richer datasets. Gigascience 2015;4:7.

28. Zheng Z, Liu S, Sidorenko J, et al. Leveraging functional genomic annotations and genome coverage to improve polygenic prediction of complex traits within and between ancestries. Nat Genet 2024;56(5):767–77.

29. Craig JE, Han X, Qassim A, et al. Multitrait analysis of glaucoma identifies new risk loci and enables polygenic prediction of disease susceptibility and progression. Nat Genet 2020;52(2):160–6.

30. James G, Witten D, Hastie T, Tibshirani R. An Introduction to Statistical Learning with Applications in R, 1st ed. New York, NY: Springer, 2009; 426 p.

31. Chen T, Guestrin C. XGBoost: A Scalable Tree Boosting System. Proceedings of the 22nd ACM SIGKDD International Conference on Knowledge Discovery and Data Mining. San Francisco, California, USA: Association for Computing Machinery, 2016.

32. Lundberg SM, Erion G, Chen H, et al. From local explanations to global understanding with explainable AI for trees. Nature Machine Intelligence 2020;2(1):56–67.

33. Gao XR, Chiariglione M, Qin K, et al. Explainable machine learning aggregates polygenic risk scores and electronic health records for Alzheimer’s disease prediction. Scientific Reports 2023;13(1):450.

34. Pedregosa F, Varoquaux Gl, Gramfort A, et al. Scikit-learn: Machine Learning in Python. J Mach Learn Res 2011;12(null):2825–30, numpages = 6.

35. DeLong ER, DeLong DM, Clarke-Pearson DL. Comparing the areas under two or more correlated receiver operating characteristic curves: a nonparametric approach. Biometrics 1988;44(3):837–45.

36. MacGregor S, Ong JS, An J, et al. Genome-wide association study of intraocular pressure uncovers new pathways to glaucoma. Nat Genet 2018;50(8):1067–71.

37. Varma R, Ying-Lai M, Francis BA, et al. Prevalence of open-angle glaucoma and ocular hypertension in Latinos: the Los Angeles Latino Eye Study. Ophthalmology 2004;111(8):1439–48.

38. MacGregor S, Ni G, Seviiri M, et al. Step change in glaucoma polygenic risk score performance enables clinical utility and disease prediction across all major ancestries. medRxiv 2026:2026.01.23.26344675.

39. Martschenko DO, Wand H, Young JL, Wojcik GL. Including multiracial individuals is crucial for race, ethnicity and ancestry frameworks in genetics and genomics. Nat Genet 2023;55(6):895–900.

40. Gao XR, Chiariglione M, Arch AJ. Whole-exome sequencing study identifies rare variants and genes associated with intraocular pressure and glaucoma. Nature Communications 2022;13(1):7376.

41. Han X, Gharahkhani P, Hamel AR, et al. Large-scale multitrait genome-wide association analyses identify hundreds of glaucoma risk loci. Nat Genet 2023;55(7):1116–25.

